# Prevalence of mental health illness in Nigeria: protocol for a systematic review and meta-analysis

**DOI:** 10.1101/2025.02.22.25322716

**Authors:** Aminu Kende Abubakar, Mohammed Nakodi Yisa, Sarah Oreoluwa Olukorode, Jolaade Musa, Oluwafemi Temitayo Oyadiran, Temitayo Rebecca Okusanya, Samuel Ogunlade, Moshood Abiodun Kuyebi, Daniel Olofin, Moshood Olanrewaju Omotayo, Ajibola Ibraheem Abioye

## Abstract

**Background:** Mental health disorders pose a major public health challenge worldwide, with one in eight individuals affected, particularly by anxiety and depression. In Nigeria, mental health issues are similarly prevalent, yet access to mental healthcare remains limited. Recent policy initiatives have increased governmental interest, but mental health research in Nigeria remains limited and fragmented.

**Methods:** This protocol outlines a systematic review and meta-analysis methodology to quantify the prevalence of mental health disorders in Nigeria. The review follows the Preferred Reporting Items for Systematic Review and Meta-Analysis Protocols (PRISMA-P) and Joanna Briggs Institute (JBI) guidelines and includes cross-sectional studies, surveys, and cohort studies providing prevalence data on mental disorders diagnosed by healthcare professionals or identified via validated screening tools in the country. Data sources include PubMed/Medline, EMBASE, and African Journals Online (AJOL), covering publications up to April 2024. Study selection, data extraction, and quality assessment will be conducted independently by multiple reviewers. A random-effects meta-analysis model will be used to synthesize data, with sensitivity and subgroup analyses to explore heterogeneity.

**Discussion:** This systematic review and meta-analysis provides an estimate of the prevalence of mental health disorders in Nigeria, informing future public health policy and resource allocation. These findings will contribute significantly to understanding mental health in Nigeria and support the development of effective strategies and policies.

Systematic review registration: This protocol is registered with PROSPERO (CRD42024559090).

## Introduction

Mental health disorders represent a significant public health challenge globally. One in every eight individuals lives with some form of mental disorder, and anxiety and depression are the most prevalent.(1) These conditions are highly prevalent in low- and middle-income countries (LMICs), such as Nigeria(2), and mental health often remains underresearched and underreported in these regions. Although the prevalence of mental health disorders in Nigeria is comparable to that in most developed countries, access to mental healthcare in Nigeria is limited.(3) A study showed that up to 12.1% of adults living in Nigerians had a lifetime rate of at least one Diagnostic and Statistical Manual (DSM-IV) disorder.(4) Additionally, 20–30% of the population is estimated to suffer from mental health problems.(5) This figure is large given Nigeria’s estimated population of more than 200 million people. Unfortunately, public awareness of mental health issues is limited, and mental health myths have persisted.(6) Recent policy initiatives such as the National Mental Health Act 2021 and the National Suicide Prevention Strategic Framework in November 2023 suggest increased interest from national health authorities. Mental health research in Nigeria remains limited and fragmented, as in other developing regions.

The emergence of fully structured diagnostic interviews that do not require highly trained clinicians for their administration has made large-scale and replicable epidemiological studies of mental disorders possible. Even with this development, such studies in developing countries are hampered by a lack of resources and are particularly rare in Africa.(7) The literature in Nigeria, which is mainly cross-sectional studies, has focused on specific regions or subgroups, including HIV patients, adolescents, elderly individuals, and incarcerated individuals. Moreover, the findings from general population studies have shown inconsistencies. Nationally representative studies are needed to guide effective planning, policymaking, and resource allocation to address mental health needs at the national level. In the meantime, systematic reviews with careful and comprehensive synthesis may allow us to quantify the burden of mental illnesses in Nigeria to the extent possible.

The primary aim of this study was to systematically review the literature to estimate the current and historical prevalence of mental disorders in Nigeria. An attempt will be made to estimate the prevalence of unique diagnoses by severity and population category. This review is structured around three key objectives:

1. to determine the overall prevalence of mental disorders among the adult population and to identify the specific types of mental disorders prevalent in Nigeria.

2. to determine the overall prevalence of mental disorders among children, along with identifying the types of disorders common in this age group.

3. to evaluate the prevalence of suicidal behaviors in the general population and explore the associated risk factors.

This protocol outlines the methodology for conducting a systematic review and meta-analysis that will quantify the prevalence of mental health disorders among the Nigerian population. It will detail the search strategies, inclusion criteria, data extraction processes, study quality assessment and statistical methods that will be used in the study.

## METHODS

This protocol will follow the publishing guidelines set forth by the PRISMA-P(8) checklist and the JBI methodology for systematic reviews on prevalence.(9) The protocol is registered with PROSPERO (CRD42024559090). Any amendments will be noted in the relevant sections of the protocol, along with the date the changes were implemented, and will be updated in PROSPERO and reported in the final article.

### Eligibility criteria

These eligibility criteria are structured according to JBI’s CoCoPop framework.(9)

### Condition

General mental morbidity or psychopathology, which includes any specific diagnosis and suicidality. Eligible studies must involve mental disorders diagnosed by healthcare professionals or identified via validated screening or diagnostic tools.

### Context

We focus on studies that provide estimates of the prevalence or proportion of mental disorders, specifically among individuals residing in Nigeria, and present data that are representative of either the general population or specific subgroups defined by clear inclusion characteristics, such as people living with HIV (PLHIV).

### Population

There will be no restrictions regarding the age, sex, or subgroup of the populations studied. Studies conducted in a psychiatric clinic or behavioral science population were excluded.

### Type of Studies

To be included, studies will be either cross-sectional surveys or studies providing baseline data from cohort studies or trials that measure the prevalence of mental disorders. These studies must be conducted within Nigeria and designed to represent the community or defined subgroups. Studies were excluded if they did not focus on the prevalence of mental disorders, were case□control studies, were systematic reviews or meta□analyses, presented only case reports, were duplicates providing no new data, involved animal or cell line research, or did not study human populations.

#### Information Sources

A systematic search of PubMed/Medline (U.S. National Library of Medicine), EMBASE (Elsevier) and AJOL will be undertaken. As we are primarily interested in the current and historical prevalence of mental disorders in Nigeria, we will examine publications from the inception of the databases to date.

#### Search strategy

The search strings will involve a combination of MeSH terms (Medical Subject Headings), Emtree terms, and topics by which articles are indexed for the PubMed/Medline and Embase databases, as well as relevant text and keywords. In addition, a manual search of the references of selected articles and related systematic reviews will also be performed to identify relevant articles that were not found through database searches.

## STUDY RECORDS

### Data management

Database records will be exported to a Zotero library for managing search results and removing duplicates, Rayyan for study selection, a custom spreadsheet for data extraction, and the National Institutes of Health (NIH) Quality Assessment Tool(10) for evaluating the methodological quality of the studies.

### Selection process

The study selection will be conducted by all the investigators across three rounds: an initial title/abstract review to exclude nonqualifying studies, a full article review of the remaining hits to determine eligibility, and, finally, data extraction from the full texts of the eligible studies. Study selection in the first two rounds will be performed in duplicate by two independent investigators, with any disagreements resolved through discussion with a third reviewer. The results of the screening and study selection process are reported in the final manuscript and presented in a PRISMA 2020 flow diagram.

### Data collection process

Data extraction will be conducted independently by assigned investigators via a standardized spreadsheet developed by A.I.A., which includes risk of bias assessments. In cases of missing or unclear study data, we contacted the study authors for clarification.

### Data Items

The data extraction form is structured into five sections to systematically collect and standardize key information from each study.

#### Study Identity

This section collects key identifiers and bibliographic details of the studies, including the study identification number, publication year, and name of the first author.

#### Study characteristics

This part of the form details the demographic and operational characteristics of the study, such as the percentage of female participants, age classifications, average age at baseline, age range criteria, and title and location of the study. It also includes a description of the study setting, primary objective, type, sampling strategy, duration, and whether it was conducted during a pandemic, along with specific details on the mental disorder studied, its diagnostic criteria, measurement methods, and assessment tools used.

#### Study Results

This section records the quantitative outcomes from the study, specifically the prevalence of mental disorders, confidence intervals, and total number of participants, along with the number of confirmed cases.

#### Quality of studies assessment

This section evaluates the risk of bias in each study by asking specific questions about the clarity of the study population definition, participation rates, selection uniformity, and representativeness of the sample, as well as whether the mental illness was consistently and accurately assessed across the study. The overall study quality was then rated as “good,” “fair,” or “poor” on the basis of these responses. The risk of bias will be evaluated via the National Institutes of Health (NIH) Quality Assessment Tool for Observational Cohorts and Cross-Sectional Studies(10), which assess both study and outcome levels. This assessment will inform data synthesis, particularly in sensitivity analyses and interpretation of findings.

#### Risk Factors

The final section documents demographic and health-related variables that might influence the prevalence of mental disorders, such as marital status, religious affiliation, educational attainment, smoking status, the prevalence of antiretroviral therapy usage, and other health conditions, such as hypertension, diabetes, body mass index, HIV status, and employment status.

## OUTCOME

The primary outcomes include general mental morbidity, which assesses a broad range of mental health disorders; specific diagnoses, which focus on the prevalence and characteristics of particular mental disorders; and suicidality, which involves evaluating the incidence of suicidal thoughts, plans, and attempts.

## DATA SYNTHESIS

A quantitative synthesis will be conducted for studies providing data on the prevalence of psychiatric disorders and associated risk factors, with inclusion criteria on the basis of outcomes measured by similar tools.

The main summary measure will be the pooled prevalence of psychiatric disorders obtained from a random-effects meta-analysis model to accommodate variability in the true prevalence measure. Separate meta-analyses will be conducted for community-dwelling adults, primary care clinic attendees, and other groups defined by demographic, occupational and clinical characteristics. Heterogeneity will be assessed with the I^2^ statistic. I^2^ will be regarded as minimal if 0–40%, moderate if >40–60%, substantial if 60–80% and considerable if >80% (11).

Sensitivity and subgroup analyses will be conducted to test the robustness of the review findings and explore sources of heterogeneity, respectively, on the basis of the screening or diagnostic approach employed, patient demographics, study setting, state or region, and other factors. We will assess publication bias via funnel plots and Egger’s test for meta-analyses with ≥10 included studies. *P* values will be 2-sided, and significance will be set at a *p* value < 0.05 for the meta-analyses.

## Data Availability

All data produced in the present study are available upon reasonable request to the authors

## CONFIDENCE IN CUMULATIVE EVIDENCE

For each pooled prevalence measure, we will assess the quality of the evidence via a modification of the GRADE approach(12). The certainty of evidence will be rated down if included studies are not representative (risk of bias), if the pooled estimate is imprecise or if there are too few studies (imprecision), if the point estimates and confidence intervals substantially differ from one another (inconsistency), or if investigators used different diagnostic approaches or tools across studies or if the diagnostic approach employed is likely to have been an inadequate surrogate (indirectness).

## DISCUSSION

This systematic review and meta-analysis protocol outlines a structured approach to examining the prevalence of mental health disorders and their associated risk factors in Nigeria. Our methodology is designed to ensure robust, reliable findings that can inform future public health policy. We aim to manage study variability and enhance the generalizability of the findings via a random effects model, as well as by conducting several different meta-analyses for different populations and clinical groups. Sensitivity and subgroup analyses explore heterogeneity sources and assess the robustness of our results. The results of this study will significantly contribute to our understanding of mental health and support improved strategies and policies.

## ABBREVIATIONS

PRIMSA-P: Preferred Reporting Items for Systematic Review and Meta-Analysis Protocols
JBI: Joanna Briggs Institute
AJOL: African Journals Online
LMICs: Low- and middle-income countries
DSM: Diagnostic and Statistical Manual
HIV: Human immunodeficiency virus
PLHIV: People living with HIV.
MeSH: Medical Subject Headings
NIH: National Institutes of Health

## DECLARATIONS

### Ethics approval and consent to participate

Not applicable

### Consent for publication

Not applicable

### Availability of data and materials

Not applicable

### Authors’ contributions

All the authors contributed from conception to writing, and all the authors have read and approved the final draft.

## Acknowledgments

None

---

**PRISMA-P (Preferred Reporting Items for Systematic review and Meta-Analysis Protocols) 2015 checklist: recommended items to address in a systematic review protocol\***

| Section and topic | Item No | Checklist item | Page where item is reported |
| --- | --- | --- | --- |
| **ADMINISTRATIVE INFORMATION** |  |  |  |
| Title: |  |  |  |
| Identification | 1a | Identify the report as a protocol of a systematic review | 1 |
| Update | 1b | If the protocol is for an update of a previous systematic review, identify as such |  |
| Registration | 2 | If registered, provide the name of the registry (such as PROSPERO) and registration number | 1 |
| Authors: |  |  |  |
| Contact | 3a | Provide name, institutional affiliation, e-mail address of all protocol authors; provide physical mailing address of corresponding author | 1 |
| Contributions | 3b | Describe contributions of protocol authors and identify the guarantor of the review | 7 |
| Amendments | 4 | If the protocol represents an amendment of a previously completed or published protocol, identify as such and list changes; otherwise, state plan for documenting important protocol amendments | 3 |
| Support: |  |  | 7 |
| Sources | 5a | Indicate sources of financial or other support for the review |  |
| Sponsor | 5b | Provide name for the review funder and/or sponsor |  |
| Role of sponsor or funder | 5c | Describe roles of funder(s), sponsor(s), and/or institution(s), if any, in developing the protocol |  |
| **INTRODUCTION** |  |  |  |
| Rationale | 6 | Describe the rationale for the review in the context of what is already known | 3 |
| Objectives | 7 | Provide an explicit statement of the question(s) the review will address with reference to participants, interventions, comparators, and outcomes (PICO) | 3 |
| **METHODS** |  |  |  |
| Eligibility criteria | 8 | Specify the study characteristics (such as PICO, study design, setting, time frame) and report characteristics (such as years considered, language, publication status) to be used as criteria for eligibility for the review | 4 |
| Information sources | 9 | Describe all intended information sources (such as electronic databases, contact with study authors, trial registers or other gray literature sources) with planned dates of coverage | 4 |
| Search strategy | 10 | Present draft of search strategy to be used for at least one electronic database, including planned limits, such that it could be repeated | 4 |
| Study records: |  |  | 4 |
| Data | 11a | Describe the mechanism(s) that will be used to manage records |  |
| management |  | and data throughout the review |  |
| Selection process | 11b | State the process that will be used for selecting studies (such as two independent reviewers) through each phase of the review (that is, screening, eligibility and inclusion in meta-analysis) | 5 |
| Data collection process | 11c | Describe planned method of extracting data from reports (such as piloting forms, done independently, in duplicate), any processes for obtaining and confirming data from investigators | 5 |
| Data items | 12 | List and define all variables for which data will be sought (such as PICO items, funding sources), any preplanned data assumptions and simplifications | 5 |
| Outcomes and prioritization | 13 | List and define all outcomes for which data will be sought, including prioritization of main and additional outcomes, with rationale | 6 |
| Risk of bias in individual studies | 14 | Describe anticipated methods for assessing risk of bias of individual studies, including whether this will be done at the outcome or study level, or both; state how this information will be used in data synthesis | 5 |
| Data synthesis | 15a | Describe criteria under which study data will be quantitatively synthesized | 6 |
| | 15b | If data are appropriate for quantitative synthesis, describe planned summary measures, methods of handling data and methods of combining data from studies, including any planned exploration of consistency (such as $I^2$, Kendall's $\tau$) | 6 |
|  | 15c | Describe any proposed additional analyses (such as sensitivity or subgroup analyses, meta-regression) | 6 |
|  | 15d | If quantitative synthesis is not appropriate, describe the type of summary planned |  |
| Meta-bias(es) | 16 | Specify any planned assessment of meta-bias(es) (such as publication bias across studies, selective reporting within studies) | 6 |
| Confidence in cumulative evidence | 17 | Describe how the strength of the body of evidence will be assessed (such as GRADE) | 6 |

